# Evaluation of a CZT-based photon-counting detector CT prototype for low-dose lung cancer screening using patient-specific lung phantoms

**DOI:** 10.64898/2025.12.30.25343218

**Authors:** Kai Mei, Leonid Roshkovan, Sandra S. Halliburton, Shobhit Sharma, Steve Ross, Zhou Yu, Richard Thompson, Leening P. Liu, Ali H. Dhanaliwala, Harold I. Litt, Peter B. Noël

**Author notes:** Corresponding Authors: Kai Mei, Address: John Morgan Building 56, 3620 Hamilton Walk, Philadelphia, PA 19104.

## Abstract

**Objectives:** To evaluate the clinical performance of a cadmium-zinc-telluride-(CZT-) based photon-counting computed tomography (PCCT) system for low-dose lung cancer screening (LCS-LDCT) using patient-specific 3D-printed lung phantoms, and to compare its image quality and radiomics consistency with a conventional energy-integrating detector CT (EIDCT) system.

**Methods:** Six 3D-printed lung phantoms, derived from patient CT datasets and representing various lesion types (solid, part-solid, and ground-glass), were imaged on PCCT and EIDCT scanners at matched dose levels (1.6 - 20.4 mGy). Quantitative image metrics, Hounsfield unit (HU) accuracy, image noise, and contrast-to-noise ratio (CNR), were assessed across dose levels. Radiomic features were extracted for each lesion and analyzed via principal component analysis to quantify feature consistency (within-cluster distance) and lesion type separability.

**Results:** PCCT demonstrated significantly lower image noise and higher CNR compared with EIDCT, particularly at lower dose levels. HU values were consistent across doses for both systems, with reduced variability in PCCT (coefficient of variation < 0.004). Radiomics analysis revealed tighter clustering (reduced within-cluster distances) and comparable lesion type separability between PCCT and EIDCT, indicating enhanced feature stability and lesion differentiation. Qualitative review confirmed superior lesion conspicuity and margin delineation with PCCT.

**Conclusions:** CZT-based PCCT outperforms conventional EIDCT in quantitative and qualitative imaging metrics for LCS-LDCT, enabling superior image quality and radiomics reproducibility at reduced radiation doses. These findings support the clinical translation of PCCT for lung cancer screening and radiomics-based lesion characterization.

**Key points:**

- PCCT offers reduced image noise and improved CNR performance, especially at ultra-low doses.
- HU stability and radiomics reproducibility are enhanced with PCCT.

**Clinical relevance statement:**

- PCCT may enable further dose reduction without compromising diagnostic accuracy in LCS-LDCT.

## 1 Introduction

Lung cancer is the most diagnosed cancer and the leading cause of cancer-related deaths worldwide, with almost 2.5 million cases and 1.8 million deaths annually [1]. Global efforts to reduce mortality from lung cancer include screening using low dose computed tomography (LDCT) in high-risk populations with the goal of identifying malignancy at an earlier stage of disease [2]. Landmark large-scale clinical trials, such as National Lung Screening Trial (NLST) [3] and Nederlands-Leuvens Longkanker Screenings Onderzoek (NELSON) [4], show lung cancer screening using LDCT (LCS-LDCT) can reduce mortality compared to no screening or screening using chest X-rays. Although LDCT typically provides high sensitivity and specificity for detection of lung cancer [5], the false-positive rate remains high [6] leading to additional imaging or invasive procedures, with risk of complication and increases burden on healthcare systems [7], [8], [9].

Photon-counting CT (PCCT) has introduced a new dimension to imaging, enabling additional clinically relevant functionalities [10], [11], [12], [13]. Early work shows the potential of PCCT to improve screening LDCT thanks to its inherent benefits of smaller pixel size and improved spatial resolution, increased soft-tissue contrast from better weighting of low-energy photons, and enhanced low-dose performance from lowered electronic noise [14], [15]. Routine clinical use of PCCT in the screening setting requires further task-specific evaluations to both establish its efficacy over conventional energy-integrating detector CT (EIDCT) and to optimize its performance for lung imaging tasks.

Standardized technical phantoms are typically used for performance evaluation of CT technologies. Although these conventional phantoms allow reproducible measurement of image quality metrics, they lack anatomical realism and do not accurately represent the complexity of pulmonary structures or lesion morphology encountered in clinical practice. This constrains their relevance for evaluating application-specific imaging tasks, such as lesion detection, classification, and size measurement in LCS-LDCT. Recent advances in 3D printing and materials science have enabled development of patient-specific CT phantoms that replicate anatomical features and tissue-equivalent CT densities [16], [17], [18]. Such models facilitate clinically meaningful assessments of CT system performance by simulating realistic imaging scenarios. Moreover, quantitative analyses derived from phantom images - such as radiomics-based feature extraction - can provide additional insight into the potential of PCCT for precise lesion characterization and improved diagnostic decision-making in lung cancer screening.

In this study, we evaluate the clinical performance of a cadmium-zinc-telluride-(CZT-) based PCCT system for LCS-LDCT compared with an EIDCT system. Using custom-designed 3D-printed patient-specific lung phantoms incorporating multiple types of pulmonary lesions, we assess both qualitative and quantitative imaging metrics relevant to early lung cancer detection.

## 2 Materials and Methods

### 2.1 Phantom design

The Institutional Review Board (IRB) approved the retrospective selection of patient CT scans for the creation of 3D-printed phantoms. Six chest CT scans from patients demonstrating different lesion types and sizes were selected to represent lung lesions observed in clinical practice. The lesions (maximum 3D diameter/mean intensity) included small part-solid (36.2 mm/-560.9 HU), large part-solid (62.8 mm/-614.6 HU), small solid (23.5 mm/-164.8 HU), large solid (43.1 mm/-94.8 HU), small pure ground-glass (22.9 mm/-747.4 HU) and large pure ground-glass (48.6 mm/-734.6 HU) as shown in **Figure 1**. All scans followed a routine non-contrast chest CT protocol, with acquisition and reconstruction parameters summarized in **Supplemental Table 1.**

**Figure 1.**
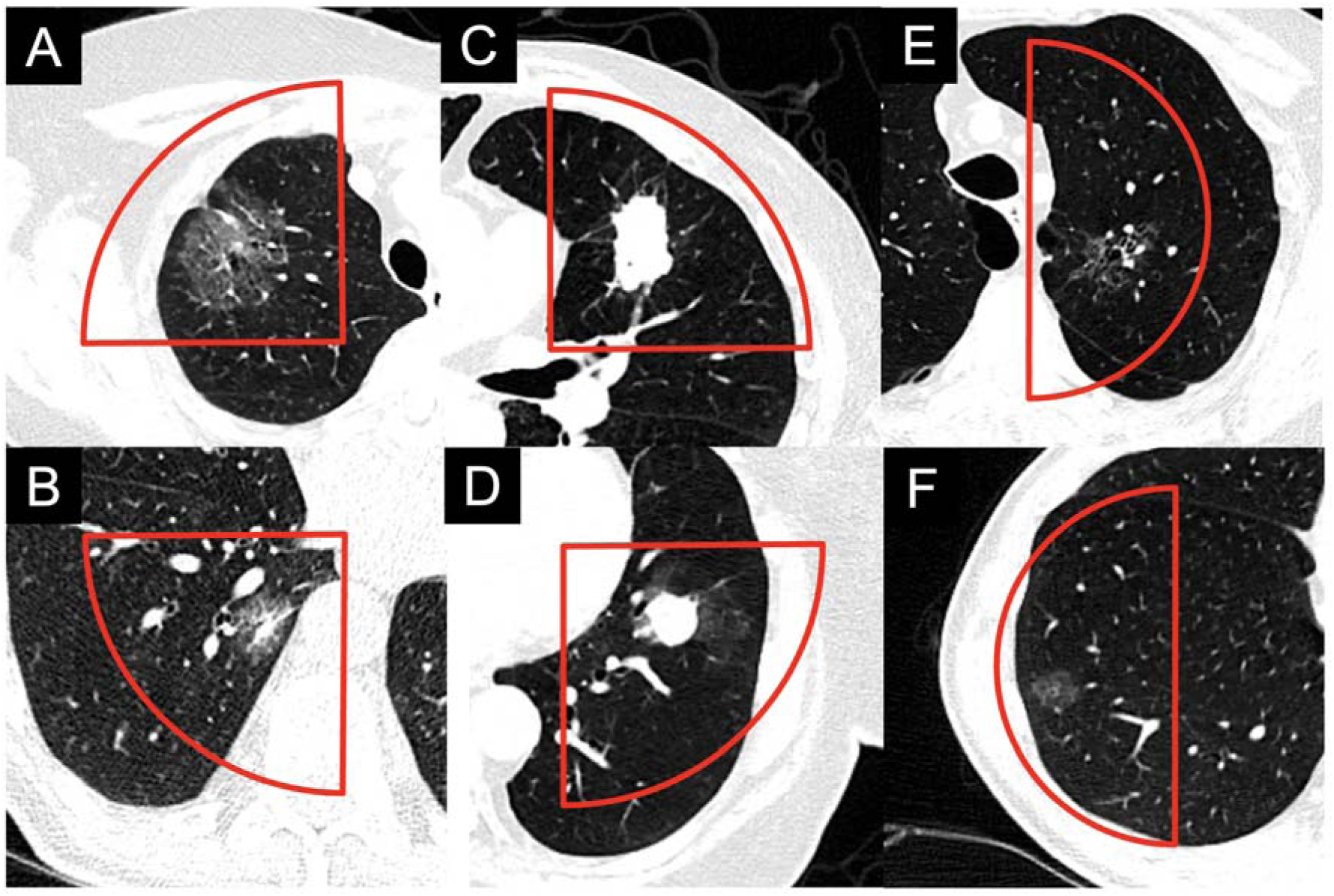
Lung lesions from patient CT scans selected for the design of two cylindrical phantoms. Phantom 1 incorporated (A) large part-solid, (B) small part-solid, (C) large solid, and (D) small solid lesions. Phantom 2 incorporated (E) large ground-glass, and (F) small ground-glass lesions. Red outline indicate the boundaries of regions containing lesions that were cropped from patient images and used to create the 3D-printed phantoms. WL/WW:-500/1000 HU.

The phantoms are cylindrical, with each lesion contained in a quarter or a half sector of the circular cross-section. Four lesions (part-solid: large and small; solid: large and small, **Figure 1A-D**) were incorporated into a Ø 200 mm x 32 mm phantom (Phantom 1)

The remaining two lesions (ground-glass: large and small, **Figure 1E-F**) were incorporated into a Ø 150 mm x 40 mm phantom (Phantom 2).

### 2.2 Fabrication of Lung Phantoms via PixelPrint-Based 3D Printing

Lung phantoms were produced using the PixelPrint technology [16], [19], [20] via fused-filament fabrication (LulzBot TAZ Sidekick, M175 tool head, 0.25 mm brass nozzle; Fargo Additive Manufacturing Equipment 3D, Fargo, ND). PLA filament (1.75 mm, MakeShaper, Keene Village Plastics, Cleveland, OH) was extruded at 190°C onto a 50°C build plate. Printing was performed with a constant volumetric feed rate (18 mm³/min) and variable print head speeds (3 - 30 mm/s) to generate line widths of 1.0 - 0.1 mm, corresponding to infill ratios of 10 - 100% with fixed 1.0 mm spacing. This dynamic infill modulation achieved voxel-specific CT attenuation as prescribed in the phantom model. Each 0.2 mm layer was printed with alternating 90° orientation. The printer operated with 500 mm/s² acceleration and 8 mm/s jerk settings to ensure geometric accuracy and layer adhesion.

### 2.3 CT scanning

The 3D-printed lung phantoms were scanned on both a clinical EIDCT (Canon Aquilion Prime, Canon Medical System USA, Tustin, CA, USA) and a research prototype CZT-based PCCT (Canon Medical Research USA, Vernon Hills, IL, USA) using dose-matched protocols. To simulate a realistic patient size, the 3D-printed phantoms were placed inside the 200 mm diameter bore of a standard CT phantom (Gammex Multi-Energy CT Phantom, Sun Nuclear, Melbourne, FL, USA. **Figure 2A**). Phantom 2 with a diameter of 150 mm was first placed inside an extension ring with a water-equivalent attenuation to increase its overall diameter to 200 mm.

**Figure 2.**
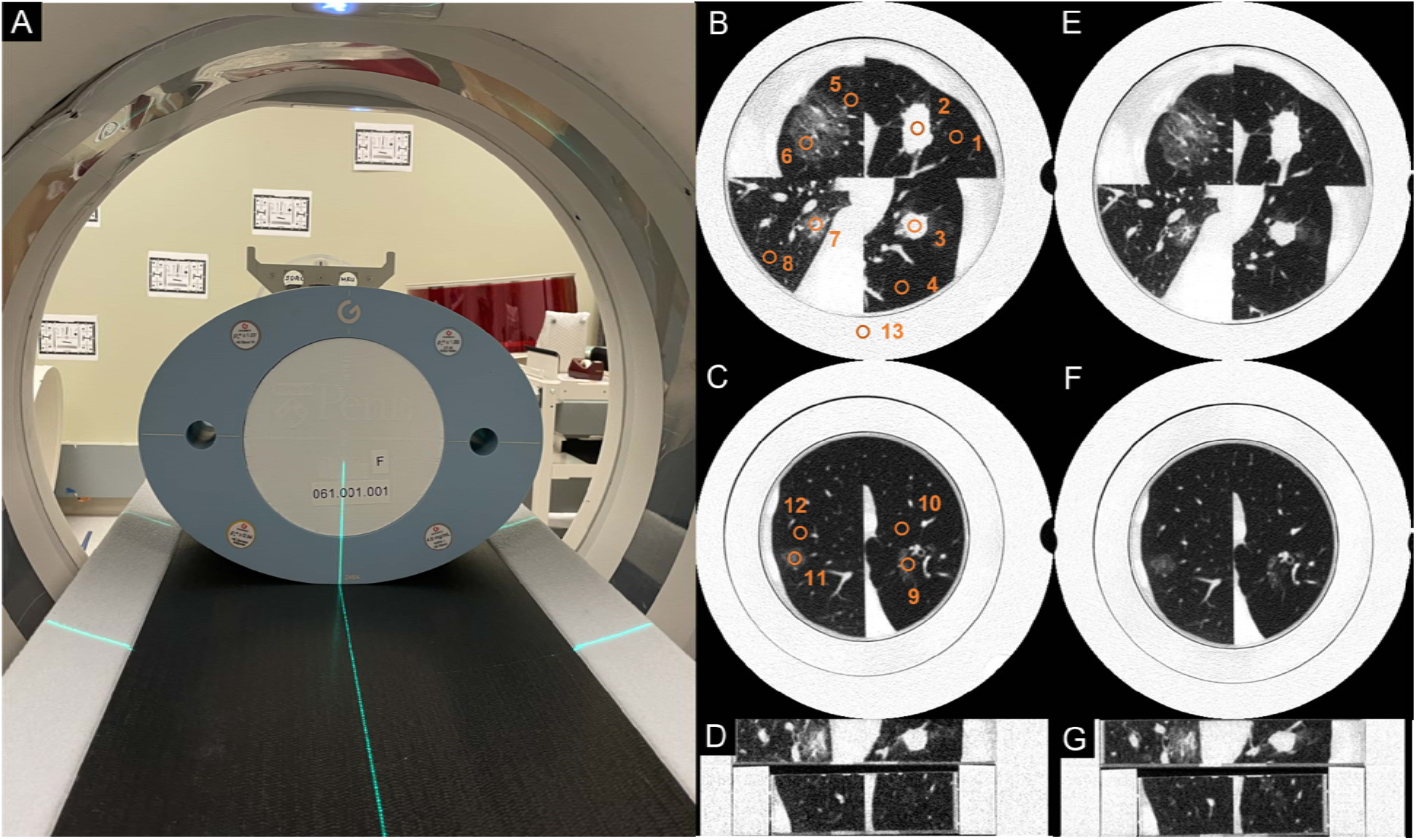
3D-printed patient-specific lung phantoms. (A) Photo showing the 3D-printed lung phantom surrounded by a Gammex MECT phantom on the patient table of a prototype CZT-based PCCT scanner. Images replicating patient lung anatomy obtained from Phantoms 1 (B) and 2 (C) using EIDCT at the highest dose (20.4 mGy) Images from Phantoms 1 (E) and (F) obtained from PCCT at highest dose (20.4 mGy). (D) and (G) are coronal view of (B/C) and (E/F). Images from EIDCT and PCCT are displayed at approximately the same location. The regions-of-interest (ROIs) used to measure mean HU, image noise, and contrast-to-noise ratios (CNRs) are marked in (B) and (C). All images are displayed using WL/WW of-500/1000 HU.

**Figure 3.**
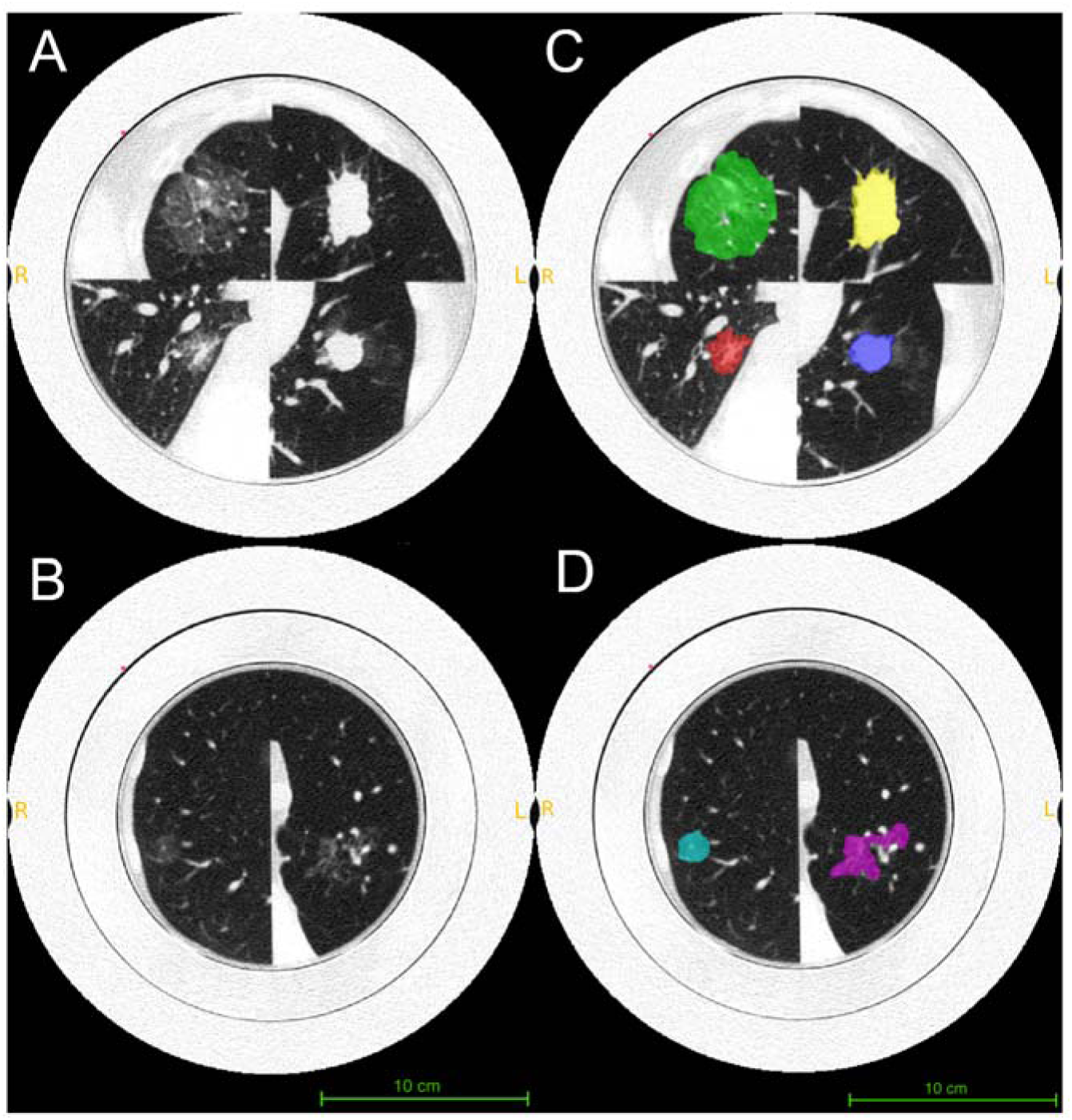
Segmentation of lesions from 3D-printed lung phantom CT images. Figures A/B show PCCT images for the 3D-printed phantoms at the highest dose (20.4 mGy). Figures C/D show the corresponding segmented lesion masks highlighted in color green (large part-solid), red (small part-solid), yellow (large solid), blue (small solid), magenta (large ground-glass), and cyan (small ground-glass). All images are displayed using WL/WW of-500/1000 HU.

For both EIDCT and PCCT systems, dose-matched scans at a tube voltage of 120 kVp were performed at CTDI_vol_ values of 20.4, 9.8, 4.9, 2.4, and 1.6 mGy. For EIDCT, this was achieved using tube current (mA) values of 250, 120, 60, 30, and 20, respectively, and a rotation time of 1s. For PCCT, the CTDI_vol_ values of 20.4, 9.8, and 4.9, were achieved using tube current values of 200, 100, and 50 mA, respectively, and rotation time of 1 s. Due to scanning constraints on the PCCT prototype, the lower CTDI_vol_ values of 2.4 and 1.6 mGy, were achieved using rotation times of 0.5 s and 0.35 s at tube current values of 50 mA. Scans at each dose were repeated three times for each scanner to estimate inter-scan variability of measurements during evaluation.

All scans were reconstructed in counting mode at normal resolution (NR) with filtered backprojection (FBP) using a lung-specific kernel (FC52) and without use of denoising methods. Images from both CT systems were reconstructed with an in-plane resolution of 0.5 mm. The nominal slice thicknesses were 0.5 mm (EIDCT) and 0.6 mm (PCCT). To facilitate direct comparison and align with standard lung image evaluation protocols, all image series were additionally generated with a uniform 1.0 mm slice thickness. A detailed list of scan and reconstruction parameters is provided in **Table 1**. Images obtained from Phantom 1 (containing four lesions) and Phantom 2 (containing 2 lesions) at the highest dose on EIDCT and PCCT systems of are shown in **Figure 2B-G**.

**Table 1.**
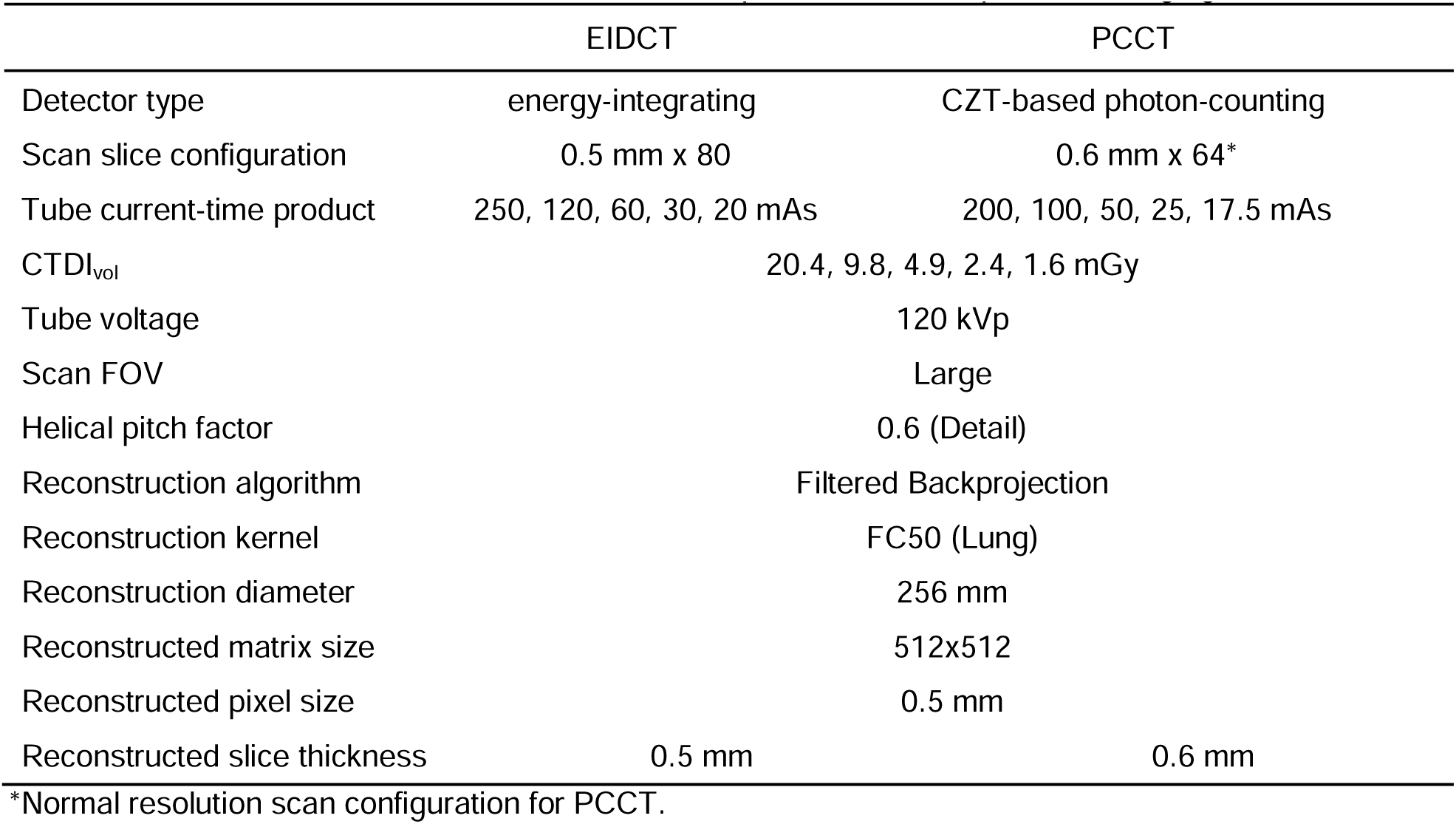
CT scan and reconstruction parameters used phantom imaging.

### 2.4 Image Evaluation

#### 2.4.1 Qualitative Evaluation

Images were assessed by an experienced radiologist (board-certified chest fellowship-trained with over 13 years of experience). Delineation of lesion margins from adjacent normal parenchyma, visualization of the ground glass component at the periphery of the lesion, and the assessment of intralesional interfaces involving lesional tissue, small bronchi, and blood vessels were considered during the review of images for all dose levels from EIDCT and PCCT.

#### 2.4.2 Attenuation, noise, and contrast-to-noise ratios

For computing quantitative image quality metrics, 13 regions-of-interest (ROIs) were placed manually on three non-overlapping central image slices. All ROIs were 8 mm in diameter and placed in homogeneous regions while avoiding high-contrast features. As shown in **Figure 2B-C**, within each slice, ROIs were placed inside all lesions and the neighboring lung parenchyma background, with another ROI placed in the solid water portion of the outer ring of the Gammex MECT phantom (water phantom). To match the locations of ROIs between systems, image registration was performed using the Random Sample Consensus (RANSAC) algorithm [21] available within the OpenCV library [22] in Python. Mean (*µ_HU_*) and standard deviation (σ*_HU_*) of HU values within each ROI were recorded. For estimating the mean (*µ_HU,i_*) and standard error (*SE_HU,i_*) of HU values associated with each material (*i* = lung parenchyma, ground-glass, part-solid, solid, or solid water), measurements across lesion sizes (large and small) and image slices were pooled. For estimating image noise (σ*_HU,lung_*), standard deviations of HU values measured for all ROIs placed within the lung parenchyma background were used. The contrast-to-noise ratios (CNRs) for solid, part-solid, and ground-glass lesions were computed using *µ_HU_* and σ*_HU_* values for ROIs placed in corresponding lesions (*k*) and the neighboring lung parenchyma background (*BG, k*) as:

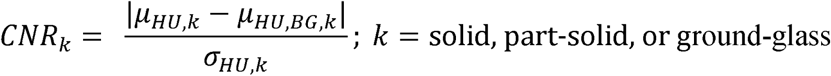

#### 2.4.3 Radiomics

All lesions were semi-automatically segmented by a radiologist using ITK-SNAP (version 4.0.2) [23]. The radiologist placed initial lesion masks on reconstructed images (window width/level = 1500/-500 HU), after which the algorithm automatically refined the mask boundaries to align with lesion edges. Manual fine-tuning was then performed to ensure that each segmentation encompassed all visible lesion components while excluding adjacent vessels traversing the nodules. For each lesion, the initial mask was created on the highest-dose images and subsequently propagated to the other dose levels with minor adjustments, minimizing the risk of excluding relevant tissue. In total, 180 lesion masks were generated (6 lesion types × 5 dose levels × 2 scanners × 3 repetitions). Radiomic feature extraction was performed in PyRadiomics [24], yielding 107 features per lesion, including 18 intensity-based, 14 shape-based, and 75 gray-level texture-based metrics.

To assess quantitatively the ability of the CT images to differentiate lesion types, Principal Component Analysis (PCA) was performed using the Statistics and Machine Learning Toolbox in MATLAB R2021b (Mathworks, Natick, MA, USA) to reduce dimensionality of the high-dimensional radiomics features. PCA reduces the dimensionality of radiomics features by transforming correlated features into a smaller set of uncorrelated principal components while ensuring the most informative variance is retained. For this study, the number of principal components used for analysis was chosen such that 95% of total variance in the original radiomics dataset was retained. Within the lower-dimensional principal component space, where points represent distinct measurements for lesions across scanner type, dose levels, and repetitions, the following metrics were computed: (1) within-cluster distance, the mean distance between points within a lesion cluster (part-solid, solid, and ground-glass) to the cluster centroid, and (2) separability, the ratio of mean pairwise distances between lesion cluster centroids and the mean of within-cluster distances for all lesion clusters. The within-cluster distance quantifies the consistency of radiomics features across dose levels and repetitions for all lesion types while separability quantifies the extent to which clusters are separated from each other on images from a given scanner.

#### 2.4.4 Statistical Analysis

All quantities are reported as mean ± standard deviation, unless stated otherwise. For testing statistical significance of differences between two groups, two-tailed paired t-test or the Wilcoxon Mann-Whitney test were performed based on the normality of the differences between the distributions as determined using the Lilliefors test. For tests involving three or more groups, one-way ANOVA was performed. All statistical tests were performed using the Statistics and Machine Learning Toolbox in MATLAB R2021b (Mathworks, Natick, MA, USA) at 5% significance level.

## 3 Results

### 3.1 Qualitative Evaluation

Qualitative evaluation by the radiologist showed that using PCCT significantly improved delineation of lesion margins from surrounding tissue. Additionally, it enhanced visualization and separation of lesional tissue from non-lesional vascular and bronchial interfaces within lesions compared to EIDCT. This improvement was observed across all doses but was especially notable for part-solid lesions at lower doses. In these cases, the differences in tissue density are often subtle and obscured by heightened noise levels associated with lower doses.

### 3.2 Attenuation, noise, and contrast-to-noise ratios

The means and standard deviations of HU values for all materials (lung parenchyma, ground-glass lesion, part-solid lesion, solid lesion, and water phantom) over all doses were-873.9 ± 5.3,-723.7 ± 9.7,-660.9 ± 2.2,-22.9 ± 2.0, and-2.9 ± 3.9 for EIDCT and - 871.5 ± 2.9,-750.1 ± 2.7,-662.8 ± 1.4,-41.8 ± 1.0,-12.7 ± 1.0 for PCCT. At each dose level, these values are summarized in **Figure 4**. The variation in measured HU values decreased with increasing dose (1.6 - 20.4 mGy) for both scanners, as indicated by the corresponding coefficients of variation (defined as the ratio of the standard deviation to the mean) of 0.006, 0.013, 0.003, 0.089, and 1.326 for EIDCT and 0.003, 0.004, 0.002, 0.024, and 0.082 for PCCT. For both EIDCT and PCCT, no statistically significant differences were found among HU values measured at different doses.

**Figure 4.**
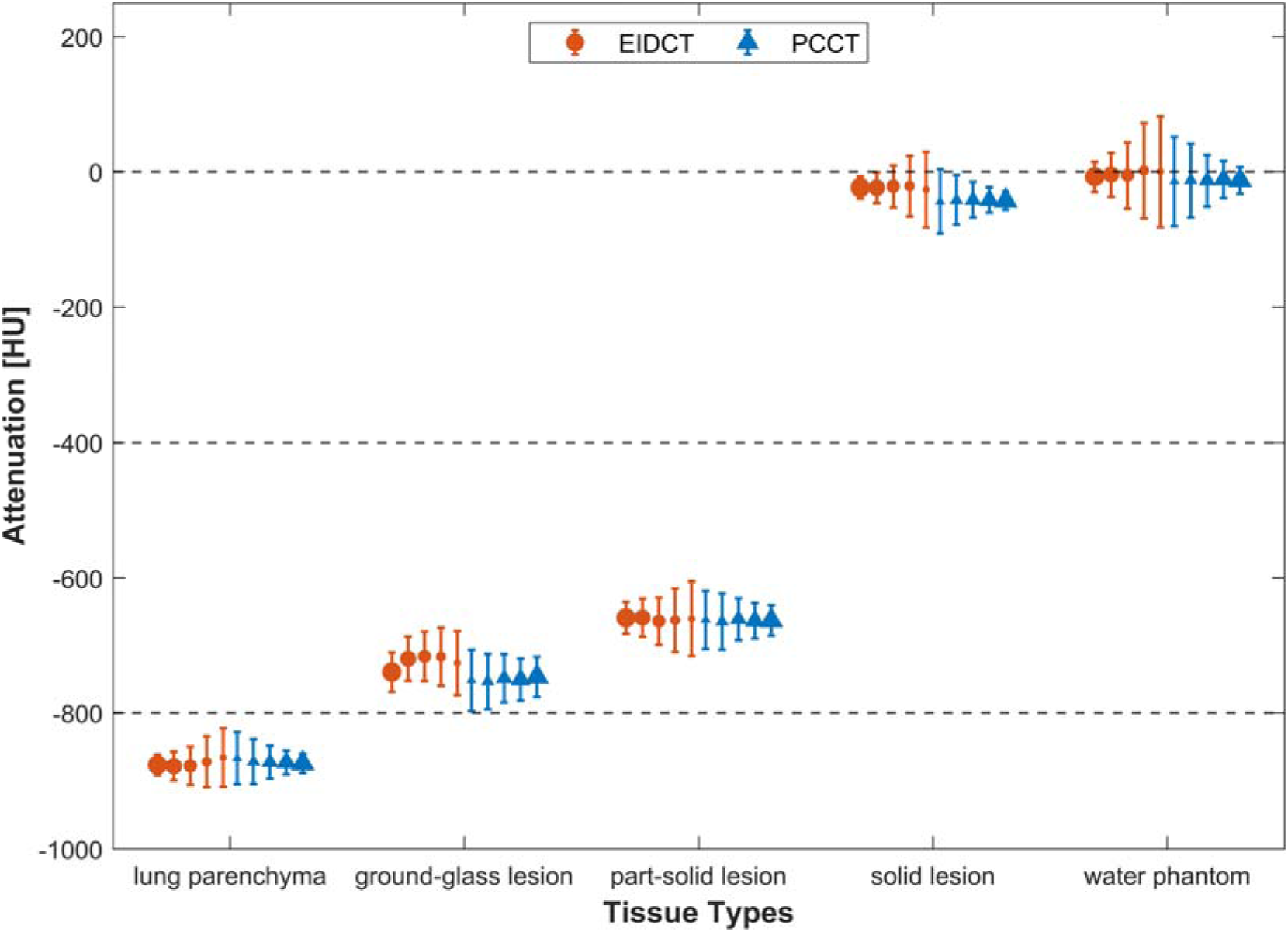
Attenuation measurements in EIDCT and PCCT images. Mean and standard deviations of HU values associated with each material of interest (lung parenchyma, ground-glass lesion, part-solid lesion, solid lesion, and water phantom). The increase in radiation dose (1.6 - 2.4 mGy) is represented by increasing marker sizes. HU values for each material at a given dose were obtained by averaging all related ROIs across lesion sizes and scan repetitions.

The distribution of image noise (σ*_HU,lung_*) for EIDCT and PCCT across all ROIs placed in lung parenchyma for all scan repetitions and doses is summarized in **Figure 5A**. Mean and standard deviation values (HU) at 1.6, 2.4, 4.9, 9.8, and 20.4 mGy were estimated to be 134.4 ± 20.9, 117.2 ± 20.1, 88.2 ± 15.1, 65.9 ± 10.8, 47.2 ± 9.2 for EIDCT and 116.9 ± 16.0, 103.5 ± 17.4, 76.8 ± 12.8, 57.9 ± 10.5, 45.8 ± 10.3 for PCCT. For both scanners, image noise decreased with increasing dose. A statistically significant reduction in noise (*p* < 0.05) was observed for PCCT over EIDCT at all doses, except at 20.4 mGy. The reductions in noise for PCCT compared to EIDCT are 3.0%, 12.2%, 12.9%, 11.6% and 13.0% for 20.4, 9.8, 4.9, 2.4 and 1.6 mGy, respectively.

**Figure 5.**
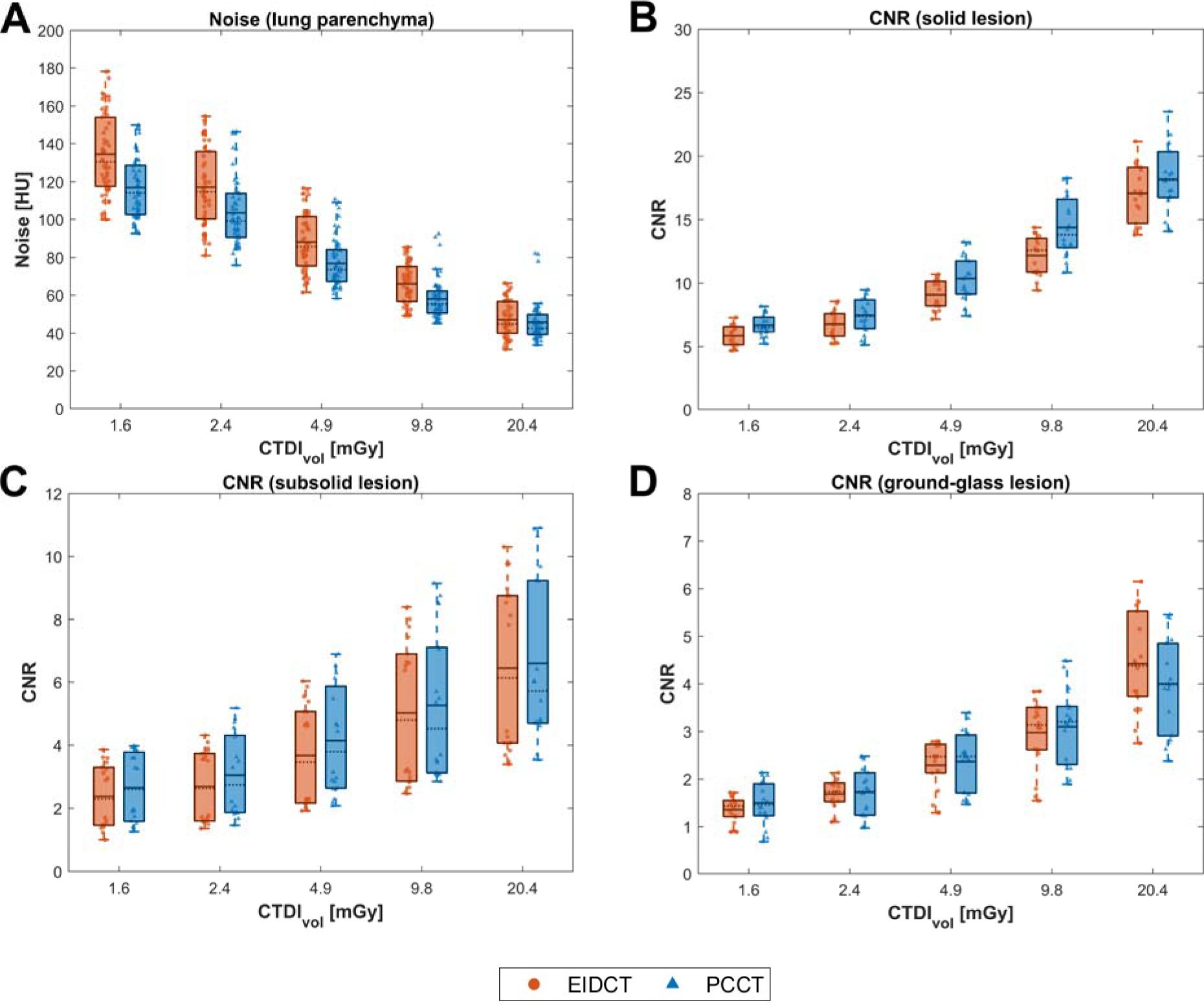
Image noise in the lung parenchyma and contrast-to-noise ratios (CNRs) in lesions in EIDCT and PCCT images. (A) Distribution of image noise for EIDCT and PCCT across all doses. (B-D) Distribution of CNR values for solid (B), part-solid (C), and ground-glass (D) lesions in EIDCT and PCCT across all dose levels. In each plot, dotted and solid lines represent the median and mean image noise values, respectively, estimated across all ROIs and scan repetitions (shown as scattered dots). The boxes indicate interquartile ranges (25th - 75th percentiles), and the whiskers extend to the most extreme values not classified as outliers. Averaging all cases at doses lower than 10 mGy, the observed overall noise from PCCT is 10.0% lower than EIDCT, and the overall CNR from PCCT is 12.4% higher.

The distribution of CNRs for solid, part-solid, and ground-glass lesions for all doses and scan repetitions is summarized for both scanners in **Figure 5B-D**. CNR values were observed to increase with increasing dose, with the highest values for solid lesions, followed by part-solid and ground-glass lesions. Average CNR for PCCT is higher than EIDCT for solid, part-solid and ground-glass type lesions. The measured CNR increases for PCCT over EIDCT are 9.0%, 10.0%, 8.7% and 12.1% at 9.8, 4.9, 2.4 and 1.6 mGy, respectively, considering all six lesions. Measured PCCT ground-glass lesion CNR was lower than EIDCT at 20.4 mGy, which can be caused by better evaluation of the true inhomogeneity inside the lesion.

A statistically significant improvement in CNR (*p* < 0.05) was observed for PCCT over EIDCT for solid lesion at all doses, except at 20.4 mGy. For part-solid lesions, CNR improved (*p* < 0.05) with PCCT except at 9.8 and 20.4 mGy. For ground-glass lesions, no significant differences in CNR were observed between PCCT and EIDCT except at 20.4 mGy where, as noted, a reduction in CNR (*p* < 0.05) was observed with PCCT.

### 3.3 Radiomics

Radiomics features for all lesions across scanner type, dose levels, and repetitions after dimensionality reduction using principal component analysis (PCA) are shown in **Figure 6**. Radiomics features in the reduced-dimensional space clustered by lesion type, showing no dependence on the scanner used for image acquisition. The number of principal components that explained 95% of total variance in the original radiomics dataset was seven. In this reduced dimensional space, the within-cluster distance for solid, part-solid, and ground-glass lesions was 5.21 ± 2.60, 5.49 ± 2.81, and 6.12 ± 2.80 for EIDCT and 5.01 ± 2.51, 4.52 ± 2.26, and 5.51 ± 2.45 for PCCT, respectively. The mean and standard deviation values of within-cluster distance were estimated across lesion sizes, scan repetitions, and radiation doses, with corresponding values of separability of 2.16 and 2.22 for EIDCT and PCCT, respectively.

**Figure 6.**
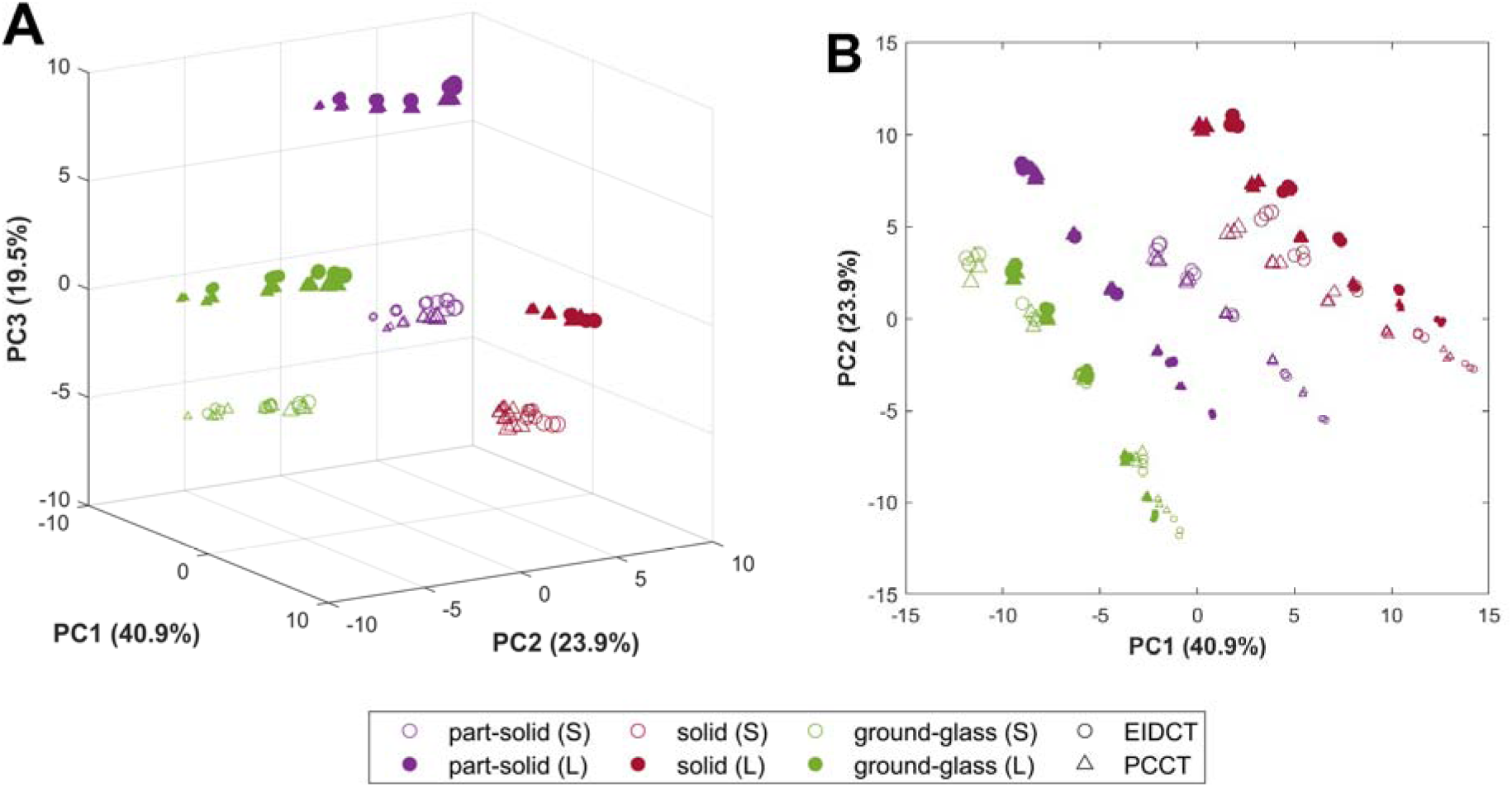
Visualization of radiomic features after dimensionality reduction using principal component analysis (PCA). 3D (A) and 2D (B) representation of radiomics features on a low-dimensional principal component space, where markers represent distinct measurements for small (S) and large (L) lesions across scanner type, dose levels, and repetitions. Principal components explaining the most variance (expressed as percentages on axes labels) in the original radiomics dataset were used for visualization. The increase in radiation dose (CTDI_vol_) is represented by increasing marker sizes.

## 4 Discussion

In this study, we used 3D-printed patient-specific lung phantoms with multiple types of lung lesions to evaluate and compare the clinical performance of CZT-based PCCT for low-dose lung cancer screening using CT. The custom phantoms, 3D-printed using PixelPrint, were based on actual patient CT datasets to provide realistic lung anatomy and represent diverse lesion morphology and size. PCCT performance was evaluated quantitatively using various conventional metrics of image quality such as HU consistency, noise, and contrast-to-noise ratios for all lesion types. Further, radiomics features were extracted for all imaged lesions and within-cluster distance and separability were computed in the reduced dimensional space to quantify the consistency of radiomics features across scans and the extent to which lesion clusters were separated from each other for each scanner.

The mean HU values for all materials (lung parenchyma, ground-glass lesion, part-solid lesion, solid lesion, and solid water from MECT phantom) on PCCT had only minimal differences from EIDCT, providing evidence that attenuation measured using the CZT-based PCCT system are comparable with conventional CT. Although there were no statistically significant differences between the HU values measured across doses (1.6 - 20.4 mGy) for both EIDCT and PCCT, the estimated coefficients of variation were much lower for PCCT, indicating better reliability of HU values. This improved reliability at lower doses is especially important for screening LDCT.

Measurements of image noise in lung parenchyma had lower mean values (3 - 13%) for PCCT compared to EIDCT across all doses, with greater improvement at lower doses. This has been observed in prior studies [12], [25] and can be attributed to improved dose efficiency due to lack of reflective septa and the effective elimination of additive electronic noise by photon-counting detectors. The reduced image noise paired with better low-energy photon weighting translated to higher CNRs for PCCT compared to EIDCT for all studied lesions: CNR was 6.4 - 18.0% higher for solid lesions, 3.1 - 14.8% for part-solid lesions, and 0 - 7.4% for ground-glass lesions. At 20.4 mGy, PCCT showed a slight (9.1%) CNR decrease for ground-glass lesions, possibly reflecting improved detection of true lesion inhomogeneity or minor artifacts. The improvements in these quantitative metrics align with subjective image quality assessments by the radiologist, which demonstrated improvement in lesion conspicuity and enhanced delineation of lesions from surrounding parenchyma for PCCT over EIDCT.

In the reduced-dimensional feature space, radiomics features clustered according to lesion type, reflecting the distinct morphological and textural characteristics of solid, part-solid, and ground-glass lesions. Within each lesion cluster, no clear dependence on scanner type was observed, suggesting that image resolution and texture are broadly comparable between EIDCT and PCCT and may partially account for the limited scanner-related differences. For all lesion types, the within-cluster distances were smaller for PCCT than EIDCT, likely due to reduced image noise and stable HU in PCCT acquisitions. This tighter clustering implies improved lesion separability with PCCT. Overall, the findings suggest that PCCT provides more reliable radiomic feature extraction and enhances differentiation among lesion types at radiation doses typical of LDCT protocols. In addition to improving differentiation between broad lesion categories, such as solid and non-solid lesions, PCCT appears to enhance the ability to distinguish lesions within the same category, indicating improved sensitivity to intra-class heterogeneity. Other recent studies of PCCT for radiomic applications indicate that although intra-scanner repeatability is high, inter-system reproducibility remains limited unless acquisition and reconstruction parameters are tightly controlled [26]. Higher spatial resolution and lower noise in PCCT improve the reliability of morphologic radiomic features [27], whereas texture features remain sensitive to reconstruction energy, kernel, and dose settings [28]. To enable robust use of PCCT in radiomics, particularly for lung nodule analysis, standardized imaging protocols and selection of energy-independent reproducible features are essential[29], [30], [31], [32], [33].

This study has some limitations. First, we studied only six lesions across three lesion types and two sizes to evaluate PCCT for LCS-LDCT. Future studies need to include a larger, more diverse sample of lesions to fully represent clinical reality and strengthen statistical conclusions. Second, we did not extensively evaluate the effect of scan and reconstruction parameters on performance. Although radiation dose was varied during acquisition, other parameters potentially impacting the studied metrics such as spatial resolution, reconstruction algorithm, and reconstruction kernel were held constant. In particular, the difference between normal resolution images as studied here and ultra-high resolution or spectral images also available from PCCT could impact results. Third, lesion segmentation performed prior to the extraction of radiomics features was achieved semi-manually by a single experienced radiologist such that any impact of segmentation variability was not captured in the results. Future studies should evaluate the effect of inter-and intra-expert variability on segmentation and its subsequent impact on radiomics quantification.

## 5 Conclusion

This study demonstrates that CZT-based PCCT offers advantages over conventional EIDCT for lung cancer screening with LDCT; with PCCT consistently outperformed EIDCT across quantitative and qualitative metrics, particularly at lower radiation doses. The findings support the clinical translation of lung cancer screening to PCCT technology, where the combination of reduced image noise and enhanced lesion conspicuity could enable further radiation dose reduction while maintaining or improving diagnostic accuracy. Further, radiomics analysis revealed more consistent feature extraction and better lesion type separability with PCCT, suggesting improved potential for automated lesion characterization. Future research should expand into larger lesion datasets and clinical validation studies to fully establish the benefits demonstrated in this phantom-based evaluation.

## Data Availability

All data produced in the present study are available upon reasonable request to the authors.

## Acknowledgement

We would like to thank Bruno Barufaldi and Vincent Dong from Department of Radiology, University of Pennsylvania, for their helpful discussion on radiomic feature analysis in this study.

We acknowledge support through the National Institutes of Health (R01EB031592, R01EB035092) and Canon Medical Research USA.

---

**Supplemental Table 1.** Scan settings for patient exams used for phantom preparation.

|  | Part-solid<br>nodule | Part-solid<br>mass | Solid nodule | Solid mass | Ground-glass<br>nodule small | Ground-glass<br>nodule large |
| --- | --- | --- | --- | --- | --- | --- |
| Tube Voltage [kVp] | 100 | 120 | 100 | 120 | 100 | 120 |
| Tube Current/ref * [mAs] | 65/90 | 124/78 | 212/108 | 130/80 | 139/108 | 122 /80 |
| CTDIvol [mGy] | 2.68 | 6.69 | 6.73 | 7 | 4.44 | 6.55 |
| Spiral pitch factor | 1.5 | 2.6 | 2 | 1.55 | 2 | 1.95 |
| Collimation [mm] | 38.4 | 57.6 | 57.6 | 57.6 | 57.6 | 57.6 |
| Kernel | Br51f | Br49d | Br49d | Br49d | Br49d | Br49d |
| Pixel spacing [mm] | 0.57 | 0.77 | 0.77 | 0.71 | 0.77 | 0.85 |
| Slice thickness [mm] | 1.0 | 1.0 | 1.0 | 1.0 | 1.0 | 1.0 |
| Field of view size [mm] | 290 | 396 | 395 | 363 | 397 | 435 |
| Matrix size | 512 | 512 | 512 | 512 | 512 | 512 |

\*Scans were tube current modulated.

